# Restrained memory CD8^+^ T cell responses favors viral persistence and elevated IgG responses in patients with severe Long COVID

**DOI:** 10.1101/2024.02.11.24302636

**Authors:** Lucie Rodriguez, Ziyang Tan, Tadepally Lakshmikanth, Jun Wang, Hugo Barcenilla, Zoe Swank, Fanglei Zuo, Hassan Abolhassani, Ana Jimena Pavlovitch-Bedzyk, Chunlin Wang, Laura Gonzalez, Constantin Habimana Mugabo, Anette Johnsson, Yang Chen, Anna James, Jaromir Mikes, Linn Kleberg, Christopher Sundling, Mikael Björnson, Malin Nygren Bonnier, Marcus Ståhlberg, Michael Runold, Sophia Björkander, Erik Melén, Isabelle Meyts, Johan Van Weyenbergh, Qian-Pan Hammarström, Mark M Davis, David R. Walt, Nils Landegren, COVID Human Genetic Effort, Alessandro Aiuti, Giorgio Casari, Jean-Laurent Casanova, Marc Jamoulle, Judith Bruchfeld, Petter Brodin

## Abstract

During the COVID-19 pandemic it was widely described that certain individuals infected by SARS-CoV-2 experience persistent disease signs and symptoms, Long COVID, which in some cases is very severe with life changing consequences. To maximize our chances of identifying the underpinnings of this illness, we have focused on 121 of the most severe cases from >1000 patients screened in specialized clinics in Sweden and Belgium. We restricted this study to subjects with objective measures of organ damage or dysfunction, >3 months following a verified, but mild-to-moderate SARS-CoV-2 infection. By performing systems-level immunological testing and comparisons to controls fully convalescent following a similar mild/moderate COVID-19 episode, we identify elevated serological responses to SARS-CoV-2 in severe Long COVID suggestive of chronic antigen stimulation. Persistent viral reservoirs have been proposed in Long COVID and using multiple orthogonal methods for detection of SARS-CoV-2 RNA and protein in plasma we identify a subset of patients with detectable antigens, but with minimal overlap across assays, and no correlation to symptoms or immune measurements. Elevated serologic responses to SARS-CoV-2 on the other hand were inversely correlated with clonally expanded memory CD8^+^ T cells, indicating that restrained clonal expansion enables viral persistence, chronic antigen exposure and elevated IgG responses, even if antigen-detection in blood is not universally possible.

## Introduction

Severe acute respiratory syndrome coronavirus 2 (SARS-CoV-2) infections cause a wide range of disease manifestations from asymptomatic or mild infection, predominantly in the young^1^, to deadly COVID-19 pneumonia associated with hyperinflammation and coagulopathy leading to multiorgan failure and death^2^. The main determinant of developing life-threatening COVID-19 pneumonia is whether a timely type-I Interferon (IFN-I) response is elicited by infected respiratory epithelial cells and plasmacytoid dendritic cells (pDC), which then limits viral replication and allows for T cells and antibodies to clear the virus. This is illustrated by life-threatening COVID-19 in individuals with inborn errors of the IFN-I activity^3^, or individuals carrying neutralizing autoantibodies to IFN-I^4^.

We have also learned that certain children develop a post-infectious condition following mild SARS-CoV-2 infection with severe hyperinflammation, skin rashes, abdominal pain and sometimes overt intestinal inflammation^5^. This Multisystem Inflammatory Syndrome in Children (MIS-C) overlaps in part with the postinfectious vasculitis syndrome, Kawasaki disease, but immunological perturbations distinguish these conditions^6^. Intriguingly, the development of MIS-C has been linked to viral persistence in the gut^7^, and perhaps repeated activation of T cells via a pathway reminiscent of superantigen-mediated T cell activation^8–11^. Prior to this observation, Cheng and colleagues reported a possible superantigen-motif within the SARS-CoV-2 spike protein that exhibited similarity to staphylococcal enterotoxin B^12^, but more recently, similar T cell skewing has also been shown in pre-pandemic samples, indicating that other pathogens, like seasonal coronaviruses, could trigger similar reactions^13^. Moreover, recessive defects of the OAS-RNAse L pathway have been shown to explain some cases of MIS-C, indicating that heightened inflammatory responses by monocytes and other mononuclear phagocytes contribute to MIS-C^14^. Genetic factors predisposing to post-infectious Long COVID are under investigation and have not been described yet. Nevertheless, it is tempting to speculate that both postinfectious MIS-C and Long COVID could involve shared mechanisms. We have proposed disease tolerance, restrained antiviral T cell responses and viral persistence as one such possible mechanism^15^. In growing children, efficient antiviral responses in the airways^16^, could allow for prioritization of limited resources towards stature growth over systemic inflammatory responses leading to mild COVID-19, but potentially also enable viral persistence and MIS-C^15^. In women of reproductive age, efficient antiviral responses limit viral replication better than in males and could allow similar prioritization of limited resources for investments into reproduction, over antiviral T cell responses and systemic inflammation, leading to mild COVID-19 in most, but also an increased likelihood for viral persistence and higher incidence of Long COVID^15^. Viral persistence has been proposed to be the key unifying feature among these postinfectious conditions, as suggested by multiple reports involving plasma samples^17–19^, tissue imaging^20^, and tissue specimens from patients^21,22^, as well as indirect evidence of continued somatic hypermutation in SARS-CoV-2 specific B cells, months after initial infection^23^.

Long COVID or post-acute COVID-19 syndrome represents a wide spectrum of diverse symptoms ranging from mild cognitive impairment and fatigue to debilitating autonomic dysregulation, microvascular dysfunction, and sometimes life-threatening tissue damage^24^. It is well established that vaccines reduce the risk but do not fully prevent the development of Long COVID^25^, and early therapy with antiviral medication following SARS-CoV-2 infection also reduces the risk of developing Long COVID^26^. These observations offer further support to the hypothesis that viral persistence is a possible cause of postinfectious Long COVID. We have formed a Long COVID subgroup within the international COVID-Human Genetic Effort Consortium (https://www.covidhge.com/), a group investigating diverse disease manifestations following SARS-CoV-2 infections by sharing and aggregating data from around the world^27^. We focus on severe cases with objective measures of organ dysfunction and damage following mild to moderate SARS-CoV-2 infections^28^.

Here we report that patients with severe Long COVID present with variable symptoms, do not cluster in relation to organs affected or immunological states. Serologic responses to SARS-CoV-2 are strongly elevated in severe Long COVID patients as compared to convalescent controls indicating persistent antigen stimulation. Direct analyses of SARS-CoV-2 RNA and protein antigens in plasma using multiple orthogonal methods reveal subsets of antigen positive individuals, but minimal overlap between assays used, suggesting that viral persistence is not uniformly detectable in plasma and that elevated serological responses is a more sensitive immunological marker of Long COVID. We also find innate immune markers which correlate with elevated IgG responses and suggest persistent inflammatory responses. The frequency of clonally expanded SARS-CoV-2 specific memory CD8^+^ T cells on the other hand are inversely correlated with elevated IgG responses in Long COVID, which add additional indirect support for the hypothesis of viral persistence, and a possible mechanism thereof in the form of restrained, rather than exhausted cytotoxic CD8^+^ T cell responses in patients with severe Long COVID.

## Results

### Identifying patients with severe Long COVID and objective measures of disease

The diagnosis of Long COVID is vaguely defined and to ensure that all patients enrolled have an organic cause of persistent symptoms, we screened >1,000 subjects at Long COVID clinics in Leuven, Belgium & Karolinska University Hospital in Stockholm, Sweden to find the most severe cases (**Fig. 1a**). Among these subjects, we only included subjects with a verified history of SARS-CoV-2 infection and mild to moderate COVID-19 course (non-hospitalized patients) to avoid overlap with the related post-intensive care syndrome which is also associated with prolonged cognitive and physiological symptoms partially overlapping with those reported in Long COVID^29^. We also limited our inclusion to subjects with objective measures of disease and organ damage which included microvascular dysfunction shown by magnetic resonance imaging (MRI) of the heart, endothelial dysfunction by pulsatile arterial tonometry (EndoPAT), autonomic dysfunction and postural orthostatic tachycardia syndrome (POTS), hyperventilation, pulmonary air trapping or reduced carbon monoxide diffusion capacity and other respiratory abnormalities which can be objectively measured by computer tomography (**Fig. 1b and Supplementary Fig. 1a**). We included 121 patients in Belgium (n=31) and Sweden (n=90), of which 87% were female and average age was 48 (min-max, 14-72 years). Numbers of symptoms were variable in each subject and did not follow any obvious grouping within the cohort (**Fig. 1b**).

**Fig 1.**
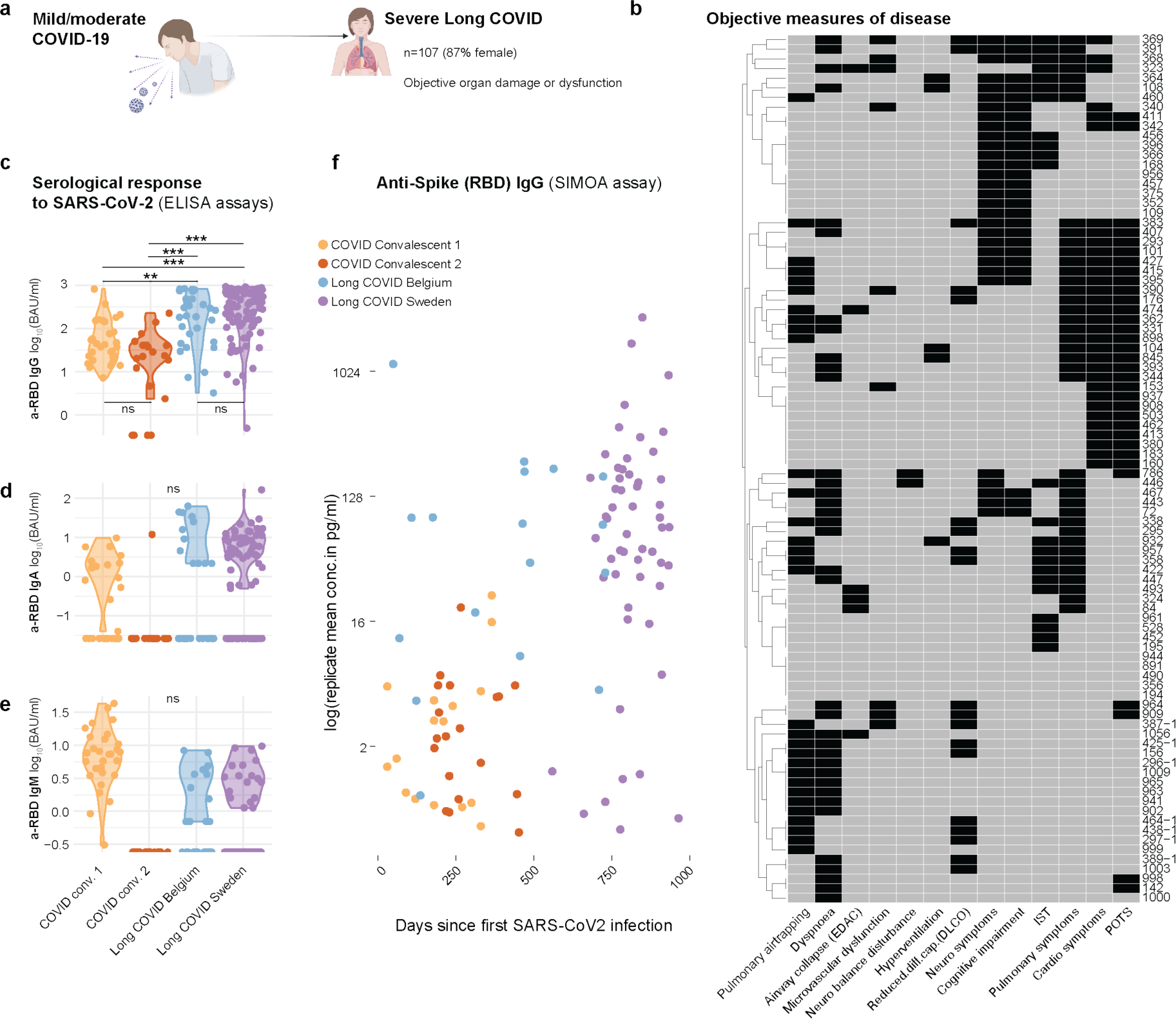
Severe Long COVID and serological responses to SARS-CoV-2. **a)** Identification of patients with severe Long COVID and objective measures of organ dysfunction/damage more than 3 months following a mild/moderate SARS-CoV-2 infection not requiring hospitalization. Cohorts from Leuven, Belgium (n=31) and Stockholm, Sweden (n=90). **b)** Distribution of objectifiable symptoms found (positive in black) among Swedish patients with Long COVID. Belgian patients shown separately in Suppl. Fig 1a. POTS: Postural orthostatic tachycardia syndrome, DLCO: Diffusion capacity of the lungs for Carbon Monoxide. EDAC: Excessive dynamic airway collapse, **c)** plasma anti-spike antibody measurements by ELISA assays surveying anti-SARS-CoV-2 spike protein receptor binding domain (RBD) IgG, **d)** IgA and **e)** IgM antibodies. BAU: Binding antibody units, **f)** Serologic analyses of anti-Spike (RBD) IgG as measured by highly sensitive SIMOA assay (Quanterix) at the indicated time point following SARS-CoV-2 infection in two cohorts of convalescent controls from Sweden and severe Long COVID cohorts from Belgium and Sweden.

### Elevated serological responses to SARS-CoV-2 in severe Long COVID

We measured antibody responses against SARS-CoV-2 using in-house ELISA assays and found that IgG responses were elevated compared to convalescent controls^30^ in both Belgian and Swedish Long COVID patients (**Fig. 1c**), however, no clear differences were observed for plasma IgA or IgM responses to SARS-CoV-2 viruses (**Fig. 1d-e**). Since standard ELISA assays are not as sensitive as single-molecule array (SIMOA) assays^31^, also for detecting SARS-CoV-2 specific IgGs^32^, we performed SIMOA assays and compared absolute IgG antibody concentrations (pg/ml) in relation to the time from first known SARS-CoV-2 infection (**Fig. 1f**). Results showed an even more pronounced difference between patients with severe Long COVID and convalescent controls, a finding that was especially clear in the Swedish cohort which included the most severe Long COVID patients (**Fig. 1f**). The result is even more striking considering that these Long COVID cases were sampled long after their acute COVID-19, after which IgG responses are expected to decline with time^23,33^, but instead showed the opposite pattern. These findings cannot be explained by SARS-CoV-2 vaccinations as there were no obvious differences regarding the timing of vaccinations or vaccination rates among the two groups.

### Persistent antigens are found in a subset of patients but with little overlap between assays used

It has been suggested that elevated serological responses could be attributed to persistent antigens and even viral reservoirs^21^. We therefore searched for persistent SARS-CoV-2 antigens in plasma samples using the SIMOA assay following reduction of endogenous SARS-CoV-2 antibodies as previously described^18^. Total spike protein was detectable in 10/100 Long COVID patients and 2/50 convalescent controls (**Fig. 2a**). The S1 subunit of spike was detectable in fewer patients as previously observed in two other cohorts^18,19^ (**Fig. 2b**), while the N antigen was detectable in 10/100 Long COVID cases, but only 1/50 convalescent controls (**Fig. 2c**). This latter finding is significant as it cannot be explained by vaccinations where the N protein is not included. These findings imply that persistent SARS-CoV-2 antigens are detectable in a subset of ∼10% of patients with severe Long COVID.

**Fig 2.**
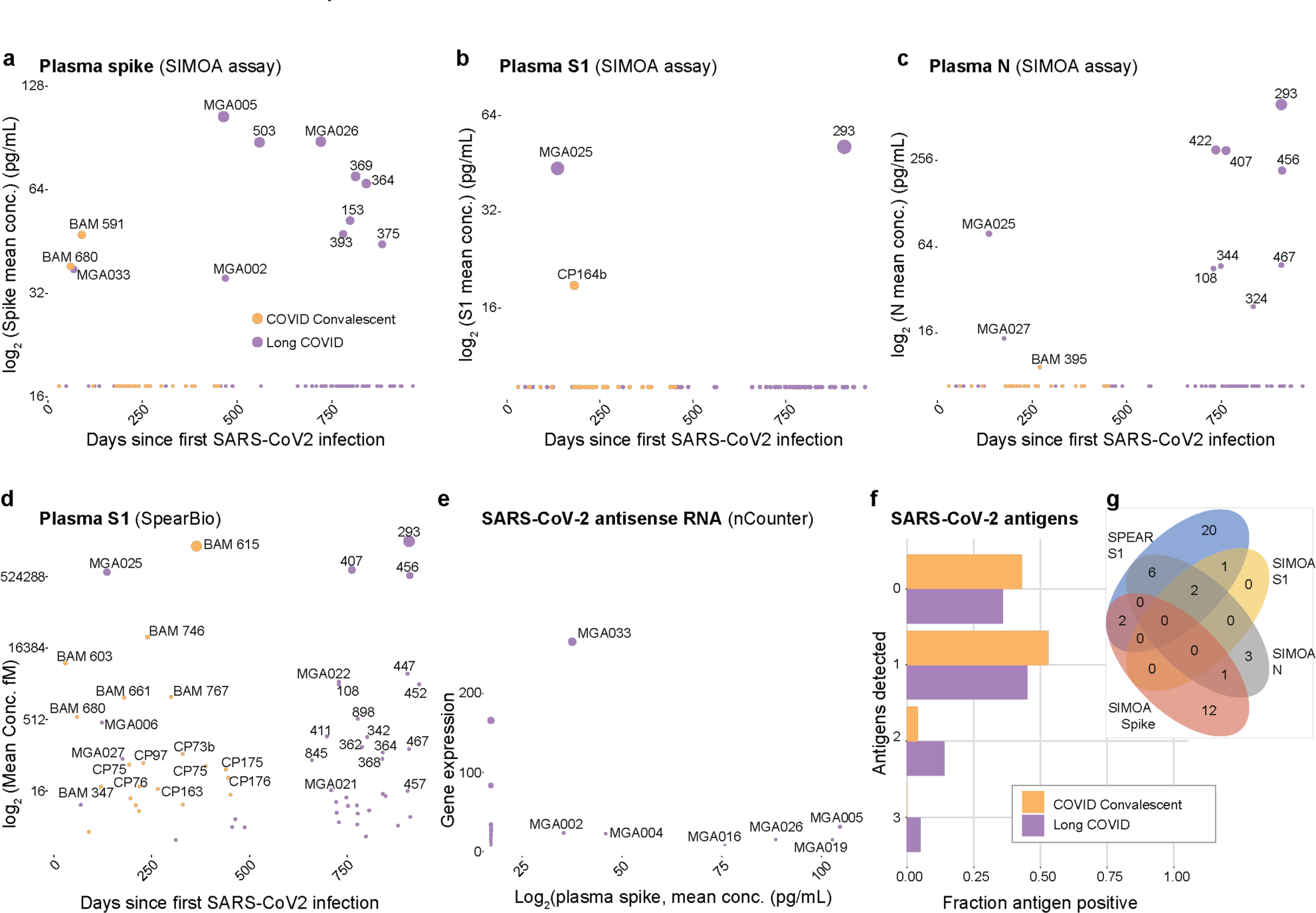
Circulating SARS-CoV-2 RNA, spike and N-protein in Long COVID patients. **a)** Plasma samples from convalescent controls (yellow, n=50) or Long COVID patients (purple, n=100) analyzed by SIMOA immunoassays for total SARS-CoV-2 spike protein, **b)** S1 subunit, or **c)** N-antigen. Numbers indicate subject ID and dot size correspond to antigen concentration, **d)** Measurement of SARS-CoV-2 S1 subunit of spike using SPEAR immunoassay (SpearBio) in convalescent controls (yellow, n=50) or Long COVID patients (purple, n=98). **e)** SARS-CoV-2 antisense RNA analyzed in (n=27) Long COVID patients from Belgian cohort and shown in relation to plasma SARS-CoV-2 spike as measured by SIMOA in the same subjects. Labels indicate subject ID and dot size by RNA expression level, **f)** Sums of SARS-CoV-2 antigen positive (protein or RNA) subjects divided by COVID convalescent (n=49) or Long COVID subjects (n=100). **g)** Venn diagram of SARS-CoV-2 protein antigen positive patients (Long COVID only).

To further investigate the detection of persistent spike protein in these same plasma samples, we also tested an orthogonal assay for S1 subunit of spike, the SPEAR immunoassay™ (SpearBio, Boston, MA). We found many more positive signals among both Long COVID (20/100) and convalescent control subjects (13/50), with no difference between these two groups (**Fig. 2d**). Both the two Long COVID patients with detectable S1 spike by SIMOA were also positive using the SPEAR immunoassay with a threshold of 16 as level of detection, while the one convalescent control CP164b was not (**Fig. 2d**). We also analyzed SARS-CoV-2 RNA using an nCounter (Nanostring technologies)^34^ in 28 of the Long COVID subjects from Belgium. Using antisense RNA from SARS-CoV-2 as a marker of ongoing viral replication we identified one subject (MGA033) positive for both antisense RNA and plasma spike by SIMOA (**Fig. 2e**). This patient was one of those sampled earliest after initial COVID-19, while other subjects with detectable spike (by SIMOA), did not show evidence of ongoing replicating virus in the form of SARS-CoV-2 antisense RNA (**Fig. 2e**). When summarizing the number of Long COVID cases and convalescent controls with any detectable antigens, we find that about half of all subjects showed some antigen-positivity, but numbers were only slightly greater in patients compared to controls (**Fig. 2f**). The presence of two or more detectable antigens was only found in Long COVID patients, representing about 10% of the cohort. There was no obvious link between detectable antigen levels in plasma and symptom groups. We also found that the different protein antigens showed limited overlap even between the two orthogonal measurements of S1 spike (**Fig. 2g**). Collectively, these results imply that either the assays used for detection of persistent antigens in plasma are not sensitive enough to detect all plasma antigens present, or that viral reservoirs confined to tissues might not leak antigens into plasma^35,36^, or that viral persistence is not a universal feature in patients with Long COVID although this latter point is at odds with the persistently elevated IgG responses to SARS-CoV-2 seen in our cohorts and in other cohorts of Long COVID patients^37^.

### Innate responses in Long COVID patients with elevated serological responses to SARS-CoV-2

Given that persistent antigen in plasma was not universal to all Long COVID cases, while elevated IgG responses to SARS-CoV-2 (SIMOA assay) more clearly distinguished patients from convalescent controls, we reasoned that elevated IgG responses might be a more sensitive biomarker to pursue further in patients with Long COVID. To understand possible immunological correlates associated with elevated SARS-CoV-2 IgG responses and possibly persistent virus, we assessed ∼26.5 million whole blood immune cells by mass cytometry using a 51-parameter panel. After clustering these cells using self-organizing maps^38^, we manually annotated the 116 clusters based on known marker combinations (**Supplementary Fig. 1b**). The cluster abundances were assessed against SARS-CoV-2 specific IgG responses measured by SIMOA as a proxy for antigen persistence. Three clusters of classical monocytes were inversely correlated with SARS-CoV-2 specific IgG (#71, #91 and #92), and one monocyte cluster (#81) expressing high CD33 was positively correlated with elevated anti-spike IgG responses (**Fig. 3a-b**). CD33-expressing classical monocytes have previously been associated with anti-tumor immune responses and favourable responses to checkpoint inhibition in lung cancer^39^, and its blockade triggers spontaneous production of TNFα, IL-1b and IL-8 suggesting a repressive function in monocytes^40^. Expansion of CD33-expressing monocytes over CD33-negative monocytes in patients with Long COVID and elevated antibody responses (**Fig. 3a-b**), indicates an attempt at dampening a persistent innate immune response to persistent antigen. To investigate the levels of cytokines and other plasma proteins we carried out Olink assays (Target inflammation™, Olink, Uppsala, Sweden) and correlated protein levels (normalized protein expression, NPX) to SARS-CoV-2 specific IgG levels. The most highly correlated proteins were circulating CD40, PD-L1, TRAIL and TNFα supporting an ongoing immune response in these patients, but not in convalescent control subjects (**Fig. 3c**). We conclude from these findings that patients with Long COVID display evidence of an ongoing innate response involving monocyte subsets and plasma proteins involved in antiviral and inflammatory responses.

**Fig 3.**
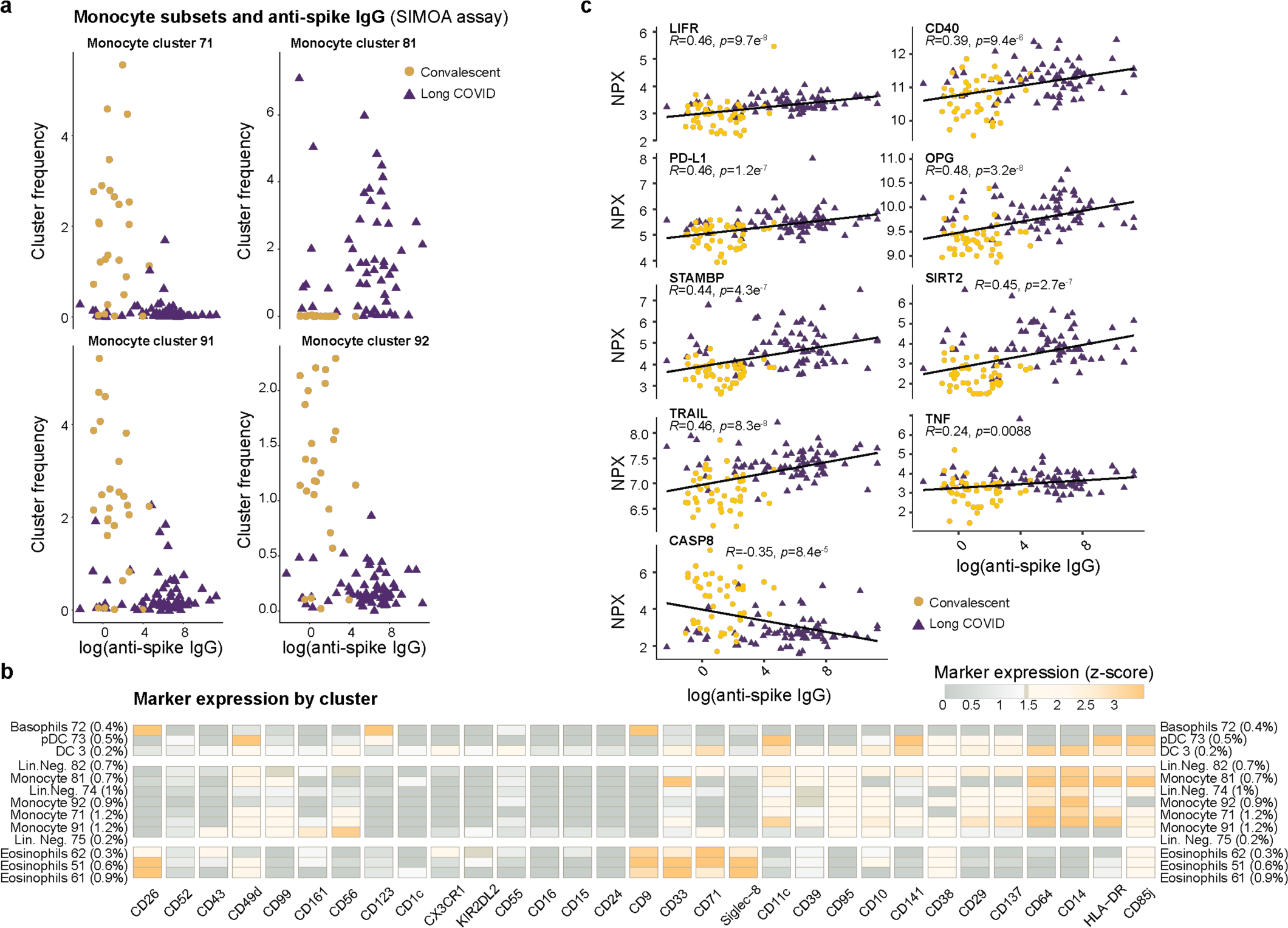
Innate immune responses in patients with severe Long COVID and variable serological responses to SARS-CoV-2. **a)** Mass cytometry profiling of whole blood samples preserved at blood draw (whole blood cell stabilizer, Cytodelics AB, Stockholm, Sweden), stained with 51-metal-conjugated probes and analyzed by Mass cytometry. FlowSOM clustering of 26,449,199 cells from 122 samples yielded 116 cell clusters, manually assigned to known cell types. Monocyte clusters 71, 81,91 and 92 differed among severe Long COVID patients (purple triangles) and COVID convalscent individuals (yellow points) and are shown in relation to serum anti-spike (RBD) IgG-antibodies (SIMOA, Quanterix). **b)** Markers expressed in the clusters shown in (a) and related myeloid cell clusters are shown colored by mean marker expression (z-score transformed), **c)** Plasma protein analyses (Olink target 96 inflammation) showing most significant protein correlates (Spearman) between anti-spike (RBD) IgG-antibodies (SIMOA, Quanterix) and normalized protein expression (NPX).

### Lack of autoantibodies targeting IFN-I in patients with severe Long COVID

Autoantibodies to type-I IFN have been associated with life-threatening COVID-19 pneumonia^4^ due to impaired IFN-I-mediated inhibition of viral replication. Such autoantibodies increase in frequency with age, are more common in males than females for unknown reason and could explain up to 20% COVID-19 deaths^41^. The reasons for the development of anti-cytokine autoantibodies are unknown in most cases, but most if not all patients with inborn errors of central tolerance due to AIRE deficiency in cis (APECED or APS1) or in trans (mutations of the alternative NF-κB pathway) all carry these autoantibodies and are highly susceptible to severe SARS-CoV-2 infections^42–44^. Since the patients with Long COVID in our cohort generally had mild to moderate acute COVID-19, we reasoned that autoantibodies to IFN-I were unlikely to be involved at the time of initial SARS-CoV-2 infection, but that de novo production of such autoantibodies following the SARS-CoV-2 infection could cause viral resurgence, persistence and Long COVID. We investigated levels of autoantibodies to four different IFNα subtypes and IL1RN but found no evidence of these autoantibodies in patients with Long COVID (**Supplementary Fig. 2**).

Lack of evidence for superantigen-mediated T cell responses in Long COVID patients. If persistent antigen, and possibly replicating viral particles are responsible for elevated anti-SARS-CoV-2 spike IgG levels and elevated markers of inflammation in patients with Long COVID, questions regarding the quality of the T cell response arise. We therefore performed single-cell TCR and mRNA-sequencing in peripheral blood mononuclear cells (PBMCs) from n=30 subjects with severe Long COVID and n=12 convalescent controls. In the postinfectious disorder MIS-C in children following SARS-CoV-2 infection, a skewing of T cell repertoires has been widely reported^8,11,45,46^. This involves the selective expansion of T cells carrying Vβ21 (TRBV11-2) paired with variable Vα genes, suggestive of a possible superantigen-mediated T cell activation and expansion. Here we find no evidence for such expansion events, either among Long COVID or convalescent controls, in either memory CD8^+^ T cells (**Fig. 4a**), or memory CD4^+^ T cells (**Fig. 4b**). This further suggests that MIS-C and Long COVID operate via distinct mechanisms.

**Fig 4.**
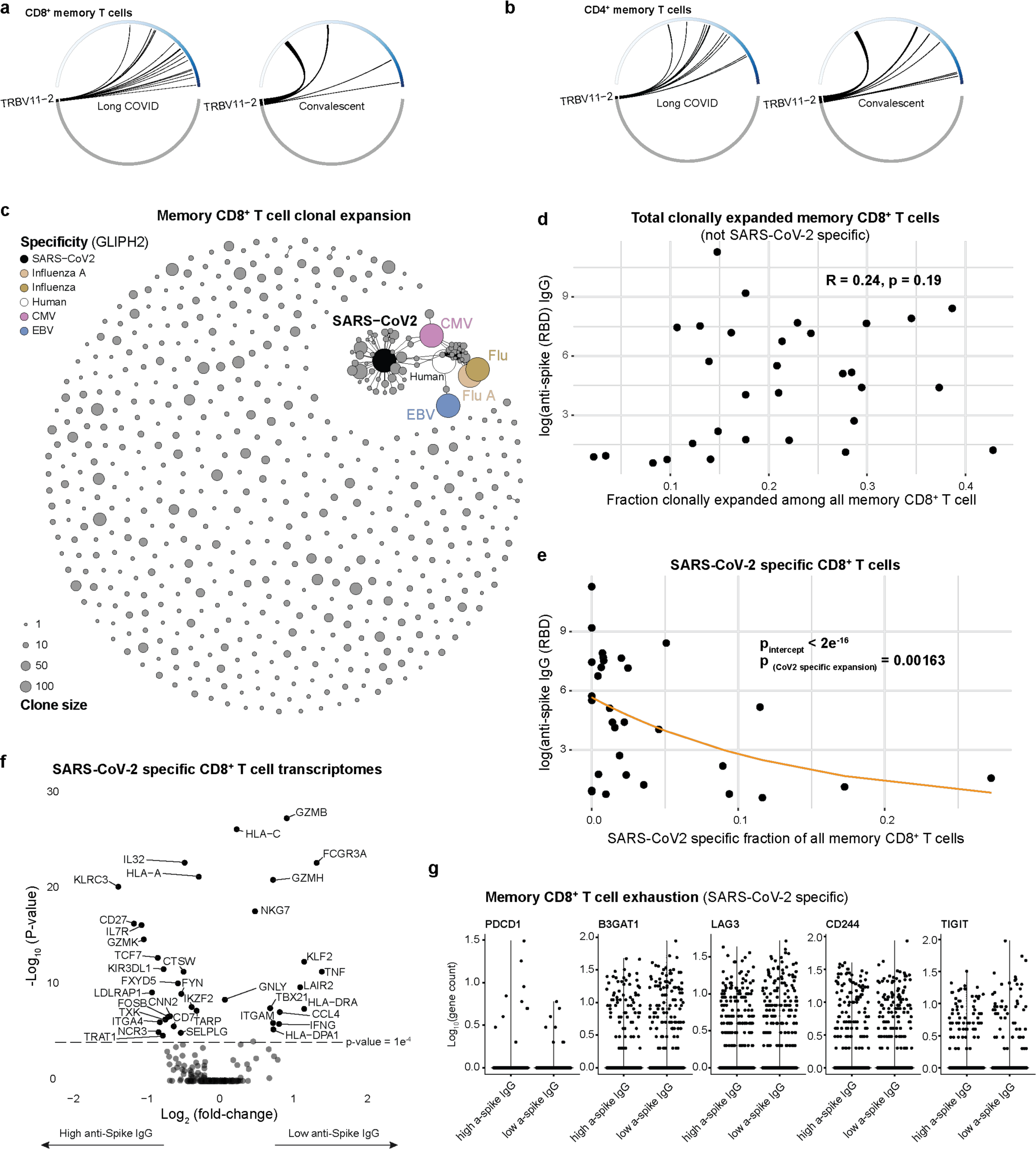
T cell responses in patients with severe Long COVID and variable serological responses to SARS-CoV-2. **a)** TRBV11-2 frequency in all (n=16,072) memory CD8^+^ T cells and **b)** memory CD4^+^ T cells (n=55,388) from 30 patients with Long COVID and 12 convalescent controls, **c)** GLIPH2 analysis showing clusters of memory CD8^+^ T cells with likely specificity for SARS-CoV-2, Flu, EBV and CMV based on multimer-sorted and sequenced T cells, **d)** Relationship between clonally expanded memory CD8^+^ T cells not specific for SARS-CoV-2 v.s. anti-SARS-CoV-2 spike (RBD) IgG measured by SIMOA assays, **e)** Relation-ship between clonally expanded and SARS-CoV-2 specific memory CD8^+^ T cells and anti-SARS-CoV-2 spike (RBD) IgG measured by SIMOA assays, **f)** Differen-tially regulated mRNA transcripts in SARS-CoV-2 specific memory CD8^+^ T cells from Long COVID patients with high and low anti-spike (RBD) IgG. **g)** mRNA transcripts associated with T cell exhaustion in SARS-CoV-2 specific memory CD8^+^ T cells from Long COVID patients with high and low anti-spike (RBD) IgG.

### SARS-CoV-2-specific T cell responses

To investigate the specificity of T cell responses is challenging with the gold standard method being multimer-tagging of antigen-specific T cells or restimulation assays. An alternative method is to infer antigen-specificity from TCR sequences, an area where important advances have been made in recent years. One such advancement is the clustering of T cell receptor Vα and Vβ sequences based on similarities in the specific amino acid residues of the Complementary Determining Region (CDR) 3 with greatest impact on peptide/MHC complex specificity^47^. Here we used this GLIPH method in its most updated version^48^ and clustered memory CD8 T cell TCR sequences (**Fig. 4c**). Based on prior data from multimer-sorted T cells with known specificities for Influenza A and B viruses, Cytomegalovirus (CMV), Epstein-Barr virus (EBV) and SARS-CoV-2 respectively^49,50^, new TCR sequence clusters obtained from Long COVID patients and convalescent controls could be mapped to these known specificities (**Fig. 4c**).

Association between SARS-CoV-2 specific T cell responses and antibody responses. The fraction of T cell clusters identified through GLIPH2 which mapped to likely SARS-CoV-2 specificity was 604 in Long COVID and 232 in convalescent controls. This expansion of T cells specific for SARS-CoV-2 in Long COVID is in line with elevated IgG responses and further suggests antigen persistence. We found no link between overall clonal expansion of memory CD8^+^ T cells and anti-spike IgG levels (**Fig. 4d**). However, when focusing on the fraction of memory CD8^+^ T cells mapping to SARS-CoV-2 by GLIPH2, an inverse relationship was found between the expansion of such cells and plasma anti-SARS-CoV-2 spike IgG levels (**Fig. 4e**). This result suggests that individuals who fail to mount a clonally expanded memory CD8^+^ T cell response to SARS-CoV-2 develop a viral reservoir with persistent antigen that drive up an elevated anti-SARS-CoV-2 spike IgG response with time. This contrast with the normal tapering of IgG titers over time following infections in convalescent individuals^23,51^.

We further characterized this memory CD8^+^ T cell response by comparing mRNA transcript levels between patients with high and low anti-SARS-CoV-2 spike IgG levels respectively. We found that individuals with lower levels of anti-spike IgG displayed a memory CD8^+^ T cell response with upregulation of molecules associated with cytotoxic Th1 responses such as GZMB, GZMH, IFNG, TNF and NKG7 (**Fig. 4f**). Patients with higher anti-spike IgG levels and possible viral persistence had a higher expression of KIR3DL1 (**Fig. 4f**), an NK cell receptor recently shown to be a marker of regulatory CD8^+^ T cells^52^. The function of these cells is to kill CD4^+^ T cells and limit autoimmune responses following infections, and accordingly, they have been found to be expanded in patients with SARS-CoV-2 infections^52^.

Moreover, a recent immunomonitoring study reported evidence of T cell exhaustion in patients with Long COVID, to a greater extent than in convalescent controls, without taking the antigen specificity of such cells into account^37^. We therefore investigated mRNA transcripts in individual memory CD8^+^ T cells from Long COVID patients with high and low levels of SARS-CoV-2-specific IgG responses respectively, but no significant differences between these subgroups were observed (**Fig. 4g**). This indicates that T cell exhaustion does not explain failure to expand productive memory CD8^+^ T cell responses to clear viral reservoirs in patients with Long COVID.

## Discussion

Our results imply that elevated SARS-CoV-2-specific IgG responses to spike (RBD), as measured by sensitive SIMOA assays, is a sensitive marker of severe Long COVID, and a likely result from chronic antigen stimulation and viral persistence. This proxy measure correlates with both innate inflammatory responses, and a restrained, unexpanded memory CD8^+^ T cell response towards SARS-CoV-2, as characterized by upregulation of negative regulators (KIR3DL1) and reduced expression of cytotoxic molecules (GZMB, GZMH), but is not associated with exhausted phenotypes.

Long COVID is a heterogeneous condition lacking clear diagnostic tests or even solid criteria, leading to a great deal of debate surrounding the condition and further complicating the interpretation of data regarding incidence and severity in populations infected by SARS-CoV-2. Here we have focused on the 121 most severe patients we have been able to identify out of >1000 cases screened in specialized Long COVID clinics in Stockholm and Leuven, all with objective measures of organ damage or dysfunction following mild to moderate SARS-CoV-2 infection^2^. By doing so, we maximize our chances of identifying the underlying biological perturbation in Long COVID and we have uncovered several immunological perturbations in these cases as compared to convalescent controls fully recovered following SARS-CoV-2 infections.

Numerous previous studies have reported the presence of SARS-CoV-2 viral particles or SARS-CoV-2 protein antigens, long after an initial infection^21^. Direct measurements of circulating SARS-CoV-2 spike proteins in the plasma of patients with Long COVID have also been reported^18,19^. Imaging studies have suggested persistent T cell activation and viral reservoirs in Long COVID^20^, and tissue biopsy studies have reported persistent viral proteins^21^ months after SARS-CoV-2 infection, but these are not always associated with persistent symptoms^23^. Antibody titers normally wane in plasma within 6 months of initial SARS-CoV-2 infection, but memory B cells can show continued somatic hypermutation suggesting viral persistence for months, even in individuals without Long COVID^23^. One study reported inverse correlation between anti-N IgG responses in the first ten days following primary SARS-CoV-2 infection and the risk of developing Long COVID^53^ and this might seem at odds with the elevated IgG responses reported herein and by others^37,54^ in patients with Long COVID cases, but this likely reflects differences between the initial and the long-term adaptive responses to SARS-CoV-2. A strong initial adaptive response might increase the chance of viral clearance and reduce the risk of Long COVID, while a sustained and elevated long-term response to SARS-CoV-2 with elevated titers occur once a viral reservoir has been established leading to chronic antigen stimulation.

Reports of innate cell perturbations in patients with Long COVID have been published but results are variable and somewhat conflicting^55^. We find monocyte redistribution in favor of CD33-expressing classical monocytes in Long COVID and elevated plasma proteins such as TNFα and TRAIL suggesting ongoing monocyte activation, in line with previous findings^56,57^. Another recent paper reported a dysregulated activation of the complement system, which could be linked to antibody-responses, either to SARS-CoV-2 or to reactivated herpesviruses^58^. Reactivation of chronic viruses has also been discussed as an initial determinant of Long COVID in patients with acute COVID-19, EBV reactivation signified patients more likely to develop persistent symptoms^59^. Accordingly, elevated levels of anti-EBV antibodies were reported in one recent study of Long COVID as compared to convalescent controls^37^. We assessed levels of IgG against EA, VCA, EBNA1 or gp42 of EBV, as well as IgM targeting gp42 and VCA but found no significant differences between Long COVID patients and convalescent controls (**Supplementary** Fig. 3). We conclude that the role of chronic viral reactivation as an underlying explanation for Long COVID remains unclear but could contribute to complement activation and microclots seen in some patients^60^.

Our finding that SARS-CoV-2 specific CD8^+^ T cells were less expanded in Long COVID cases at a frequency that was inversely correlated with the elevated serologic response, suggests a restrained antiviral T cell response as a key aspect of Long COVID pathology. Others have suggested an exhausted T cell responses in Long COVID based on elevated PD-1 expression and other marker changes^37,61^ but in our analyses we find no difference in exhaustion markers in relation to IgG responses, suggesting that the failure in viral clearance arises earlier as a consequence of a restrained antiviral CD8^+^ T cell response permitting the establishment of a viral reservoir and the development of Long COVID. It is interesting to speculate about the reasons for why predominantly females of reproductive age and children are at risk of developing postinfectious disorders (MIS-C and Long COVID) associated with SARS-CoV-2 persistence. One already known common feature of children and women of reproductive age is the very efficient IFN-I responses during acute SARS CoV-2 infection^15^. In mice the timing and source of IFN-I can either suppress or potentiate an antiviral CD8^+^ T cell response^62^ and one possible mechanisms thereof is balanced IFN-I mediated activation of STAT1 over STAT4 where the former is restraining CD8^+^ T cell responses and the latter potentiates them following viral infections^63^. Ongoing work in our laboratories are looking for possible genetic causes underlying the failure to clear SARS-Cov2 viruses and development of Long COVID^27^. All in all, our results suggest that a persistent viral reservoir is likely contributing to Long COVID pathology and ongoing efforts (NCT05823896) to remove such persisting viruses with prolonged antiviral therapies are important and hold clinical promise for delivering a much-needed cure to the community of Long COVID sufferers worldwide.

## Data Availability

All the code to reproduce figures is in our dedicated GitHub study repository: https://github.com/Brodinlab/LongCOVID_Sweden_Belgium. This also includes processed data required for analyses while raw cytometry data is uploaded to FlowRepository.org: https://flowrepository.org/id/FR-FCM-Z76S.

## Acknowledgement

We thank all the patients for participating, colleagues around the world within the COVID-HGE network, all the members of the Brodin lab and clinical teams at Karolinska University Hospital, Stockholm and in Leuven, for insightful discussions and careful monitoring of patients. This work is supported by the HORIZON HLTH-2021-DISEASE-04 program under grant agreement 01057100 (UNDINE), the Swedish research council (2021-06529) and Knut & Alice Wallenberg Foundation (VC-2021-0026). The Laboratory of Human Genetics of Infectious Diseases is supported by the Howard Hughes Medical Institute, the Rockefeller University, the St. Giles Foundation, the Fisher Center for Alzheimer’s Research Foundation, the Meyer Foundation, the JPB Foundation, the French National Research Agency (ANR) under the “Investments for the Future” program (ANR-10-IAHU-01), the Integrative Biology of Emerging Infectious Diseases Laboratory of Excellence (ANR-10-LABX-62-IBEID), the French Foundation for Medical Research (FRM) (EQU201903007798), the ANRS-COV05, ANR GENVIR (ANR-20-CE93-003), the European Union’s Horizon 2020 research and innovation programme under grant agreement No 824110 (EASI-genomics), the HORIZON-HLTH-2021-DISEASE-04 program under grant agreement 01057100 (UNDINE), the ANR-RHU COVIFERON Program (ANR-21-RHUS-08), the Square Foundation, *Grandir - Fonds de solidarité pour l’enfance*, the *Fondation du Souffle*, the SCOR Corporate Foundation for Science, Battersea & Bowery Advisory Group, William E. Ford, *General Atlantic’s* Chairman and Chief Executive Officer, Gabriel Caillaux, General Atlantic’s Co-President, Managing Director and Head of business in EMEA, and the General Atlantic Foundation, the French Ministry of Higher Education, Research, and Innovation (MESRI-COVID-19), Institut National de la Santé et de la Recherche Médicale (INSERM), REACTing-INSERM and the University of Paris Cité.

## Declaration of Interests

P.B, J.M and T.L are founders and shareholders of Cytodelics AB developing blood stabilizer solutions used in this study. P.B is an executive board member of Kancera AB (Stockholm, Sweden). D.W has a financial interest in Quanterix Corporation, a company that develops an ultra-sensitive digital immunoassay platform. He is an inventor of the SIMOA technology, a founder of the company, and also serves on its Board of Directors. Dr. Walt’s interests were reviewed and are managed by Brigham and Women’s Hospital and Partners Healthcare in accordance with their conflict-of-interest policies.

## Supplementary material

Three figures, supplementary methods, Key resource table and supplementary table with COVID-Human Genetic Effort consortium author list.

---

**Supplementary Figure 1.**
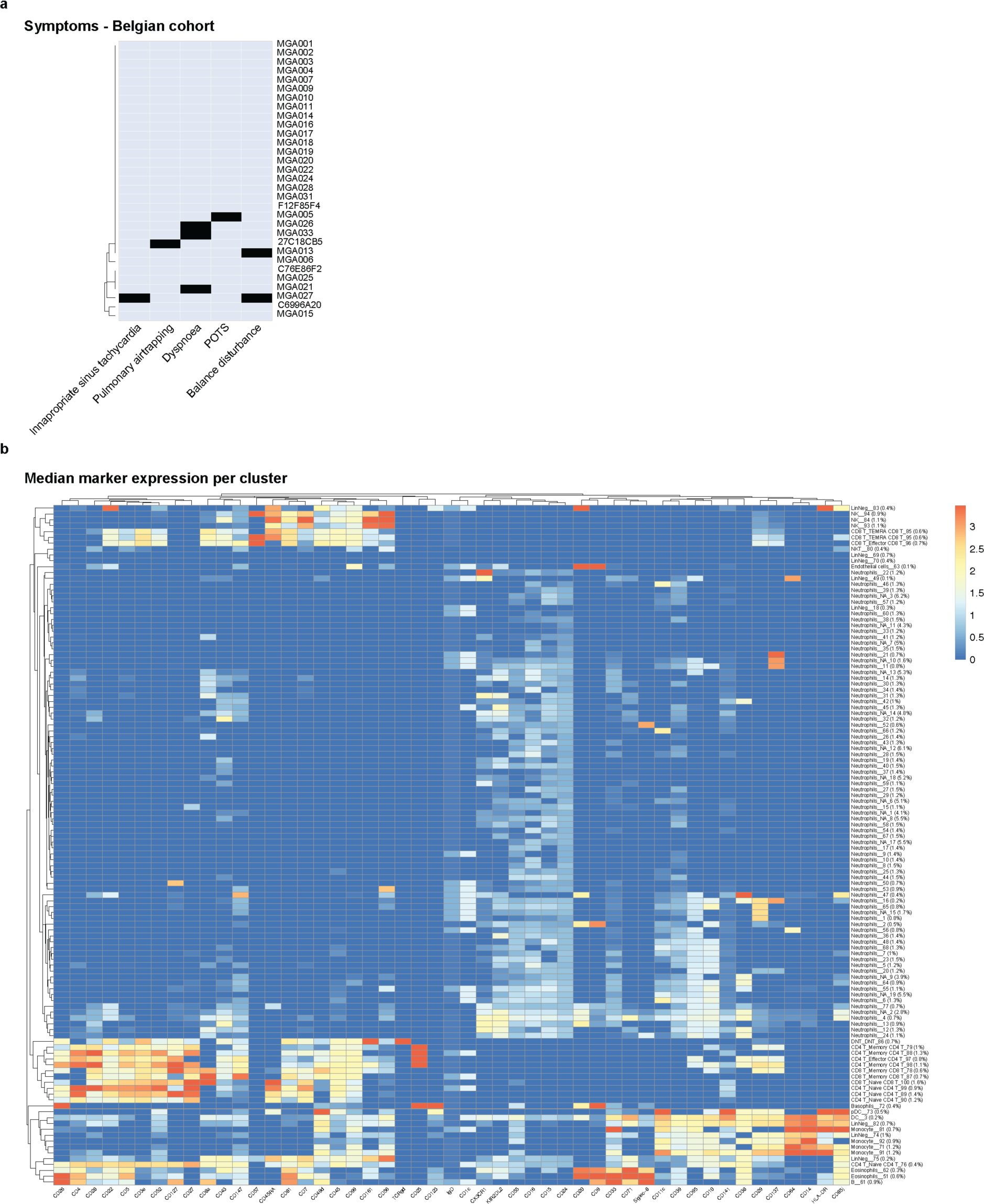
**a)** Symptoms and objective measures of diseases in Belgian cohort, **b)** Median marker expression (Z-score transformed) colored per cluster manually annotated based on known phenotypes. Cluster abundance shown as % of all cells.

**Supplementary Figure 2.**
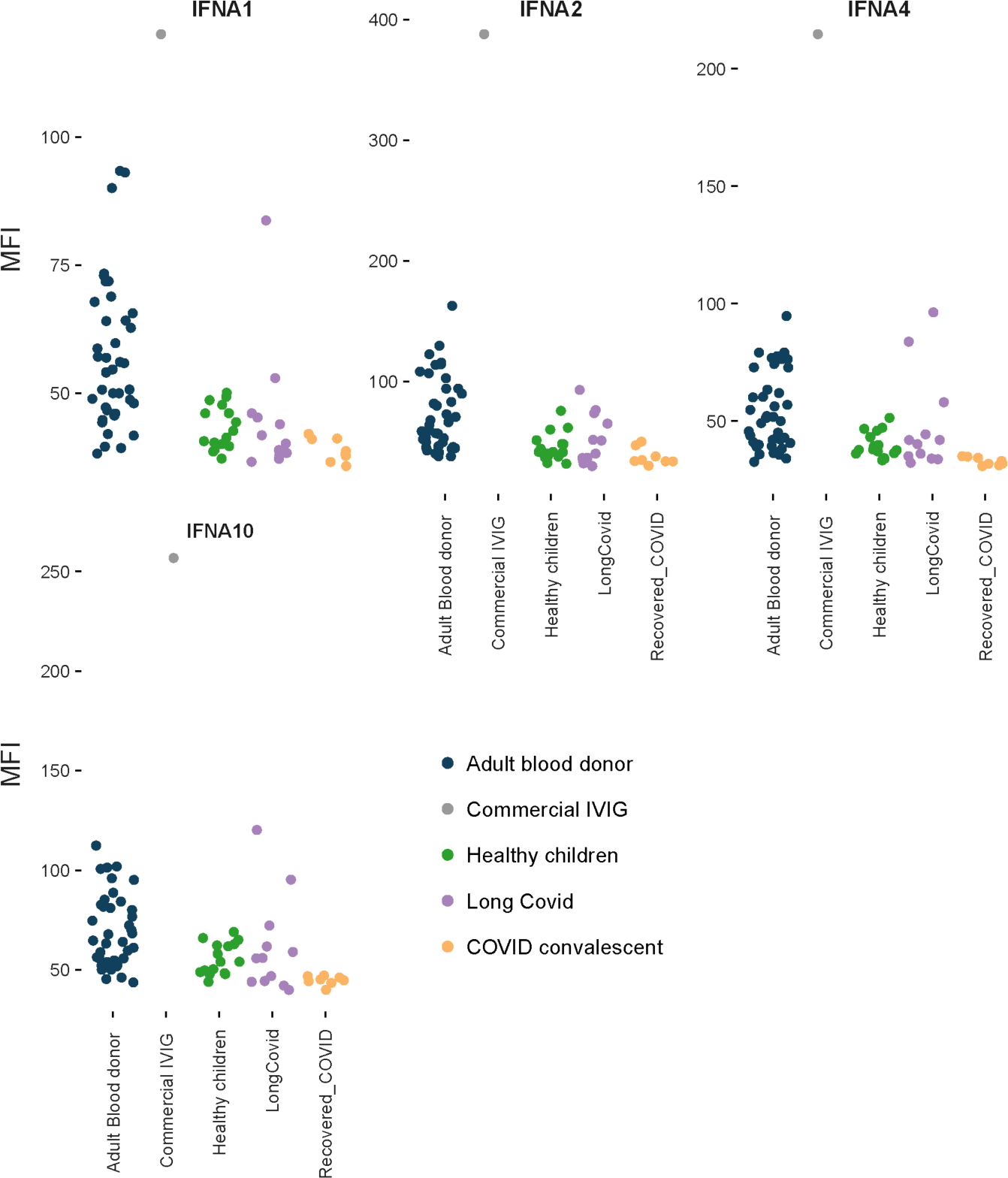
Anti-cytokine autoantibody profiling. Autoantibodies towards type-I IFN measured by xMAP flourescent bead assays from the indicated group of patients.

**Fig S3.**
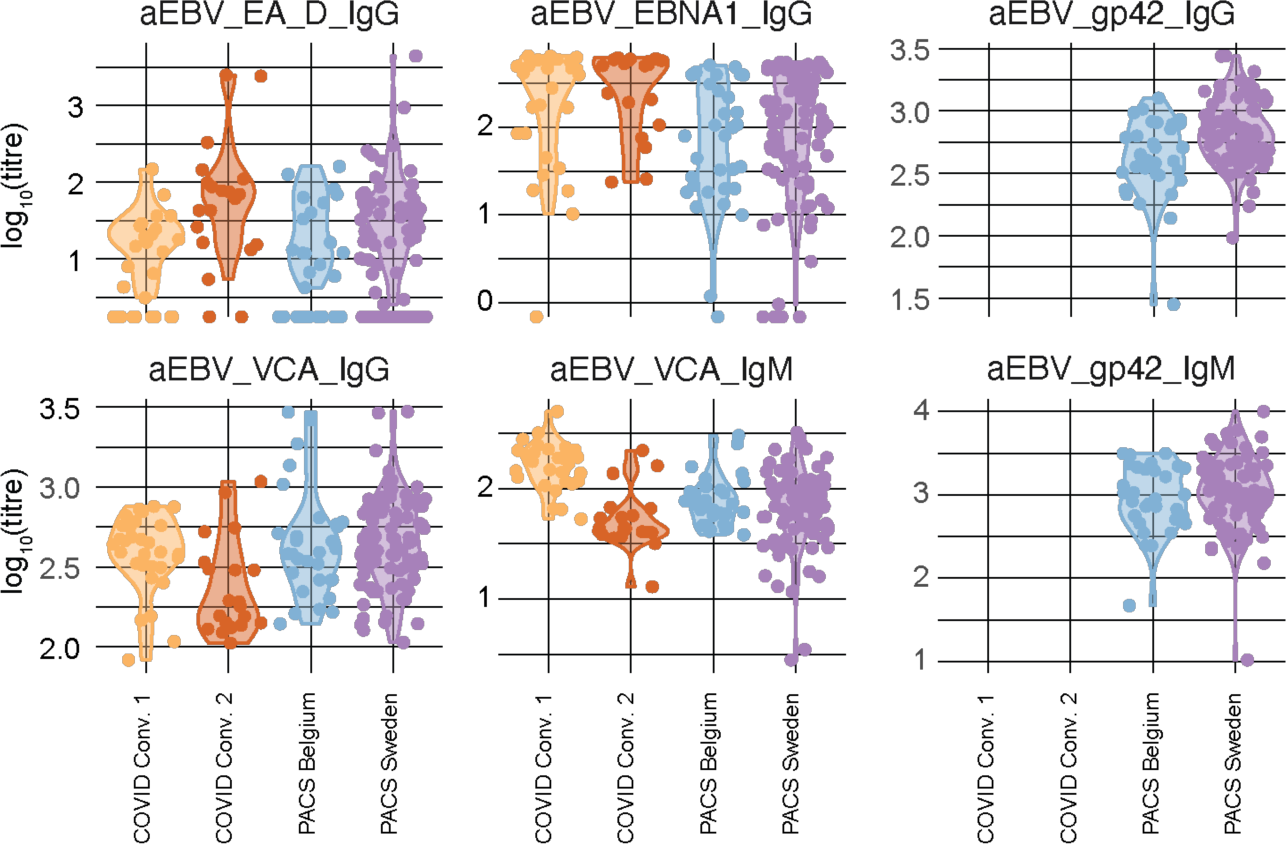
EBV serostatus in severe LongCOVID. xMAP flourescent bead assay testing the indicated antibody species binding to the indicated EBV antigen.

## Notes

### Funding Statement

This work is supported by the HORIZON HLTH-2021-DISEASE-04 program under grant agreement 01057100 (UNDINE) and the Swedish research council (2021-06529) and Knut & Alice Wallenberg Foundation (VC-2021- 0026). The Laboratory of Human Genetics of Infectious Diseases is supported by the Howard Hughes Medical Institute and the Rockefeller University and St. Giles Foundation and the Fisher Center for Alzheimers Research Foundation and the Meyer Foundation and the JPB Foundation. Also the French National Research Agency (ANR) under the Investments for the Future program (ANR-10-IAHU-01) and the Integrative Biology of Emerging Infectious Diseases Laboratory of Excellence (ANR-10-LABX-62-IBEID) and the French Foundation for Medical Research (FRM) (EQU201903007798) and the ANRS-COV05 and ANR GENVIR (ANR-20-CE93-003). Also the European Unions Horizon 2020 research and innovation programme under grant agreement No 824110 (EASI-genomics) and the ANR-RHU COVIFERON Program (ANR-21-RHUS-08) and the Square Foundation and Grandir - Fonds de solidarite pour lenfance and the Fondation du Souffle and the SCOR Corporate Foundation for Science and Battersea & Bowery Advisory Group and William E. Ford General Atlantics Chairman and Chief Executive Officer and Gabriel Caillaux and the General Atlantics Co-President and Managing Director and Head of business in EMEA and the General Atlantic Foundation and the French Ministry of Higher Education and Research and Innovation (MESRI-COVID-19). Institut National de la Sante et de la Recherche Medicale (INSERM) and REACTing-INSERM and the University of Paris Cite.

### Author Declarations

The study was approved by Swedish Ethical Review Authority (Dnr 2021-03293).

