## Supplementary material for "Restrained memory CD8^+^ T cell responses favors viral persistence and elevated IgG responses in patients with severe Long COVID"

### *METHODS*

#### *Study participants*

Following screening of >1000 subjects at the post-COVID clinics in Leuven, Belgium & Karolinska University Hospital in Stockholm, Sweden, only subjects with verified SARS-CoV-2 infection and mild to moderate COVID-19 (non-hospitalized) were included following informed consent. The study was approved by Swedish Ethical Review Authority (Dnr 2021-03293). Further, the inclusion of subjects was limited to those with objective measures of disease and organ damage which included microvascular dysfunction, postural orthostatic tachycardia syndrome (POTS), hyperventilation, air trapping or other respiratory abnormalities which could be objectively measured by physiological tests, imaging such as computer tomography or MRI. In total, 121 subjects in Belgium and Sweden were included, of which 87% were female with an average age of 48 (min 14-max 72).

#### *Whole blood sample processing*

Blood was drawn into $K_2$EDTA-containing sterile vacutainer tubes from each patient and prepped as follows: 0.5ml of blood was mixed with an equal amount of stabilizer (Cytodelics AB), incubated for 10 min at ambient temperature and stored at -80$^o$ C; 0.4ml of blood was mixed with PAXgene solution (BD Biosciences), incubated for 2 hours at ambient temp and stored at -80$^o$C; the remaining blood was centrifuged at 4$^o$C and 2000g for 10 min, following which plasma was collected into multiple aliquots and stored at -80$^o$ C.

#### *Bulk RNA sequencing of whole blood samples*

To examine changes in gene expression, RNAseq was performed using RNA extracted from blood PAXgene samples. RNA samples were prepared using a QIAcube with PAXgene blood RNA kit (Qiagen). Before cDNA library preparation, RNA quality was determined by quantifying RNA integrity number (RIN) using the Agilent 2100 bioanalyzer with the RNA 6000 Pico Kit. RNA concentration was measured by Qubit Fluorometer using the Qubit dsDNA HS Kit (Thermo Fisher Scientific). For final sequencing and cDNA library preparation, the Advanta RNA-Seq XT NGS Library Preparation Kit was used with the Juno™ system (Standard BioTools Inc.). Bulk RNA sequencing was run on a NovaSeq 6000 instrument using one flow cell SP-200 (Illumina), paired-end reads and a 2x100 read length.

***Immune cell profiling by mass cytometry***

Blood samples were mixed with a stabilizer[1] (Whole blood processing kit component; Cytodelics AB) within the first hour post blood-draw and cryopreserved according to the manufacturer's recommendations. Samples were then thawed, fixed and lysed using Lysis and Wash buffers (Whole blood processing kit; Cytodelics AB). Following fixation/lysis, 1-3x$10^6$ cells/sample were plated and cryopreserved using CRYO#20 (Cytodelics AB). For multiplexing and staining - cells were thawed at 37°C, barcoded using an automated liquid handling robotic system (Agilent technologies)[2] using the Cell-ID 20-plex Barcoding kit (Standard BioTools Inc.) as per the manufacturer's recommendations, and stained batch-wise after pooling. Briefly, cells were washed using CSB (cell staining buffer) (Standard BioTools Inc.), FcR blocked using an in-house-prepared blocking solution for 12 min at ambient temperature, following which cells were stained using a cocktail of metal conjugated antibodies targeting surface antigens (Table 1) and incubated for 30 min at 4°C. Cells were washed twice with CSB and fixed overnight using 2% formaldehyde in PBS (VWR). For acquisition by CyTOF XT, cells were stained with DNA intercalator (0.125 mM Iridium-191/193 or MaxPar Intercalator-Ir (Standard BioTools Inc.) in 2% formaldehyde and incubated for 20 min at ambient temperature. Cells were washed twice with CSB followed by two washes with Maxpar® Cell Acquisition Solution (CAS) Plus (Standard BioTools Inc.) before being filtered through a 35mm nylon mesh, diluted to 500,000 cells/ml using CAS Plus, and divided into polypropylene tubes. A total of 2x$10^6$ cells/tube in pelleted form were then placed in the chilled carousel of the CyTOF XT instrument (Standard

BioTools Inc.). EQ™ Six (EQ6) element calibration beads (Standard BioTools Inc.) were added to a tube and placed in the carousel. The autosampler of the CyTOF XT dispensed CAS Plus to the pelleted sample tubes, mixed with EQ beads 0.1X, and then acquired on CyTOF XT mass cytometers at a rate of 300-500 cells/sec using CyTOF software v8.0 with noise reduction, event length limits of 10-150 pushes, and a flow rate of 0.030 ml/min.

***Mass cytometry antibodies and reagents***

Purified antibodies were obtained in carrier/protein-free buffer and coupled to lanthanide metals using the MaxPar X8 or MCP9 antibody conjugation kits (Standard BioTools Inc.) as per the manufacturer's recommendations. Metal-conjugated antibodies were also purchased from Standard Biotools. The antibodies used for this study are listed in **Table 1.**

***Plasma protein profiling by Olink***

Plasma protein data was generated using by Olink assay, a proximity extension assay (Olink AB, Uppsala)[3]. 20uL of plasma from each sample was thawed and analysed using a Target™ Inflammation panel (Olink AB), at the Affinity Proteomics-Stockholm unit, Science for Life Laboratory, Stockholm, Sweden. In these assays, plasma proteins are dually recognized by pairs of antibodies coupled to a cDNA-strand that ligates when brought into proximity by its target, extended by a polymerase and detected using a Biomark HD 96.96 dynamic PCR array (Standard BioTools Inc.).

***Memory T cell selection from PBMCs***

Initially, Human cryopreserved PBMC samples are thawed in a 37 degree water bath, once nearly thawed,  1mL warm complete RPMI medium with benzonase (RPMI + 10% FBS + 1% P/S + 25U/ml Benzonase) were added, then transferred each PBMC sample to a 15mL centrifuge tube with around 9 mL same medium and centrifuged . The cell pellets are then resuspended in warmed RPMI medium without benzonase and cell count and viability are determined, the PBMCs are washed and centrifuged again, and a single-cell suspension of lymphocytes is prepared in cold Robosep buffer. The isolation method for untouched memory T cells were developed by using magnetic depletion of naive T cells, B cells, NK cells, monocytes, DCs, granulocytes, platelets, and erythroid cells from Human PBMC samples. PBMCs are incubated with a cocktail of biotinylated CD45RA, CD14, CD15, CD16, CD19, CD56, CD34, CD36,CD51/61, CD123, CD141, and CD235a (glycophorin A) antibodies. These cells are subsequently magnetically labelled with Anti-Biotin MicroBeads for depletion. Then the untouched memory T cell can be Isolated by using MS column from Miltenyi. The sorted cells are used for subsequent analysis, such as RNA extraction for memory T cells and DNA extraction for non-memory T cells, along with viability and cell number testing. Further experiments are performed based on the cell types obtained. Antibodies listed in **Table 2**.

***Targeted Transcriptome and TCR by BD Rhapsody for Single Cell Sequencing***

Here we use BD Rhapsody for analyzing paired chain information of TCR and targeted mRNA at single cell level, Isolated memory T cells from different human PBMC samples were resuspended in cold BD Sample Buffer, and then pooled the required number of cells from each sample to get approximately 30,000 cells for each cartridge. After loading cells to cartridges and incubated at room temperature. Cell Capture Beads were prepared and then loaded onto the cartridge. According to the BD Rhapsody protocol, cartridges were washed, cells were loaded and lysed, and cell capture beads were retrieved and washed prior to performing reverse transcription, template switch reaction and treatment with Exonuclease I, cDNA Libraries for TCR and target transcripts were continually amplified and indexed using PCR reaction. The amplification serves to enrich TCR sequences within the sample, enabling the subsequent profiling of T-cell diversity, The amplified sequences are tagged with unique identifiers, allowing for the tracking of individual TCRs. Quality of final libraries was assessed by using Agilent Bioanalyzer and quantified using a Qubit Fluorometer.

Final libraries were diluted to 2nM for paired end (150bp) sequencing on a NovaSeq sequencer (Illumina).

***Detection of auto-Abs against cytokines by multiplex beads-array***

AnteoTech Activation Kit for Multiplex Microspheres (Cat# A- LMPAKMM-10) was applied in accordance with the protocol of the manufacturer, including the optional blocking, to couple Magnetic beads (MagPlex®, Luminex Corp.) to the following commercial proteins (using 1,5 * 10^6 beads and 3 micrograms of each respective protein – one micrograms less in the cases of DSG1 and OR4K13) : CYP21A2, ADH7, ALOX15, alpha 2 macroglobulin like, CASK, Desmocolin 2, DPYSL5, Desmoglein-3, KLHL31, KLHL40, KLHL41, Laminin gamma 1, LRRC32, PADI3, PKP1, PNMA5, RARS2, SERPINB3, SERPINB4, SFN, TFAP2B, TGM1, TGM3, TMPRSS11D, TROVE2, UGP2, UNC45B, ZNF300, IFNA2, IFNA1, IFNA7, IFNA14, IFNB1, IFNE, IFNW1, IL23, Antithrombin III, Factor V, Protein S, SARS-CoV-2 Nucleocapsid, SARS-CoV-2 S protein spike, SARS-CoV-2 S protein RBD, Desmoglein-1, IFNA5, IL1F6, Prothrombin, IL22, OR4K13, IFNA6, IL1RN, JUP, IL28a, IFNA10, IFNA8, SPCS2, IL28b, IFNA16, IFNG, ADH5, IL29, IFNA17, IFNK, anti-IgG, IL6, IFNA2, IFNL4, EBNA1, ADAMS13, IFNA21, IL17A, RBM38, CXCL4, IFNA4, IL17F, ATP4A, ISG15, IFNG, IFNW1, IL17F, IL22, ApoH GM-CSF, MUSK, TNF- alpha, Protein C, IL12. Samples were diluted 1:25 in PBS prior to 1:10 dilution in Assay buffer (0,05 PBST, 3% BSA, 5% Milk). Stock of magnetic beads were sonicated for 1 minute before distribution and then mixed with storage buffer from the activation kit. Diluted samples were centrifuged 1 min at 3000 rpm, and 45 microliters of each was subsequently incubated 2 hours with 5 microliters from the stock bead solution on a shaker at 650 rpm protected from light at room temperature. Beads were then washed (3xPBS-T 0.05%) after a 2000 rpm pulse and resuspended in 50 microliters of 0.2% PFA per well followed by careful vortex. After 10 minutes of incubation at room temperature and 2000 rpm pulse, beads were washed (3xPBS-T 0.05%) and incubated with secondary antibody (Invitrogen, H10104 lot#2384336) 30 minutes at room temperature. Finally, the above washing routine was repeated, and beads were dispensed in PBS-T 0.05% before the analysis.

**Detection of antibodies specific to SARS-CoV-2 RBD**

To evaluate anti-RBD binding activity, high-binding Corning half-area plates (Corning #3690) were coated with G614 RBD (1.7 µg/ml) in PBS overnight at 4°C. Subsequently, serial dilutions of serum in PBS with 0.1% BSA were added, and the plates were incubated for 1.5 hours at room temperature. Afterward, the plates underwent a washing step and were incubated for 1 hour with horseradish peroxidase (HRP)-conjugated goat anti-human IgM (Invitrogen #A18835), goat anti-human IgG (Invitrogen #A18805), or goat anti-human IgA (Jackson #109-036-011) antibodies, all diluted 1:15000 with 0.1% BSA-PBS. The antibodies bound to the plates were visualized using tetramethylbenzidine substrate (Sigma #T0440). The color reaction was stopped with 0.5 M H2SO4 after a 10-minute incubation, and the absorbance was measured at 450 nm using an ELISA plate reader (Varioskan, Thermo Scientific). Each sample was tested in duplicate, and the mean OD values were utilized to calculate the concentration of specific antibodies.

Serum IgA and IgM antibody levels are reported as arbitrary units (AU)/ml, established based on a standard curve generated using data derived from a serially diluted highly positive in-house serum pool. IgG levels were expressed as binding antibody units (BAU)/ml after calibration with in-house standards against the WHO International Standard for anti-SARS-CoV-2 Ig (NIBSC, 20/136). The cut-off values for antibody positivity were determined through receiver operating characteristic curves, using data from convalescent COVID-19 patients and negative historical control samples. The positivity cut-offs were set at > 8.4 AU/ml for anti-RBD IgM, > 14.8 AU/ml for anti-RBD IgG, and > 0.08 AU/ml for anti-RBD IgA, achieving specificities of 96% for IgM, 97% for IgG, and 99% for IgA[4].

**Detection of Antibodies Specific to EBV Antigens**

Specific IgG and/or IgM antibodies against EBV VCA, EA-D, EBNA-1 (JENA Bioscience, Germany) and EBV (HHV-4) gH/gL/gp42 Recombinant Protein (Mybiosource, USA) were detected using ELISA, following the same procedure as described above. All antigens were coated at a concentration of 0.5 μg/ml. Specific IgG and/or IgM antibody levels are reported as AU/ml, established based on a standard curve generated using data from a serially diluted highly positive in-house serum pool. The positivity cut-offs were set at >80 AU/ml for anti-VCA IgG, >5 AU/ml for anti-EBNA1 IgG, indicating EBV seropositivity. The reactivation of latent EBV infection in EBV seropositive cases was defined as either a positive reaction with anti-VCA IgM (with cut-offs set at >181 AU/ml) or positive results with anti-EA-D IgG (with cut-offs set at >140 AU/ml).

#### *Quantification of anti-spike IgG by Simoa*

Anti-spike IgG were quantified in plasma and in supernatants of 150 samples including convalescent controls and Long COVID patient PBMCs by Simoa digital ELISA (Quanterix) with HomeBrew assays. During processing, 3 samples were flagged due to the analytical LOD of 0.015 pg/mL.

#### *Olink Preprocessing*

Plasma protein data was batch corrected and normalized based on NPX values across batches with available bridge samples using Olink's bridge normalization function in R with *OlinkAnalyze*. Plasma proteins with low variance were filtered out (>40% below LOD), with 93 immune plasma proteins remaining.

#### *Mass cytometry Preprocessing*

All unrandomized FCS files processed using CyTOF software (version 6.0.626) were directly transferred without any additional preprocessing. Data analysis was performed in R and arcsin h transformed with cofactor of 5 using the flowCore package. Beads and dead cells were filtered out. Batches were combined, and batch effects in marker expression were removed using the package sva. The resulting matrix was utilized for immune composition analysis using the FlowSOM package[5]. Briefly, we identified 30 initial clusters, annotated neutrophil clusters, and performed clustering of the remaining non-neutrophil cells with 100 total clusters. A total of 116 unique clusters were annotated manually based on median marker expression using the pheatmap package through the utilization of a z-score heatmap. A total of 15327108 cells pertaining to the 166 samples of which 133 samples were used for further analysis to solely compare PASC patients and convalescent controls.

***Single-cell TCR sequencing analysis***

Raw sequencing files were first processed by pipeline provided by BD and then analyzed with Seurat package and homemade R script. In brief, the single cell RNA expression counts were transformed by SCTransform function using regularized negative binomial regression. Afterwards, PCA was performed on the gene expression profile and the first 30 components were used for UMAP embedding. The k-nearest-neighborhood graph (k=25) was constructed and then we used Leiden algorithm to find clusters. In the determination of T cell subpopulation, cells with TRGV and TRDV genes were labelled as $\gamma\delta$T cells according to VDJ sequencing results. Then, the expressions of CD3E, CD4 and CD8A were used to label the $CD4^+$ T cells, $CD8^+$ T cells and non-T cells. In the analysis of VDJ sequences, only single cells with paired CDR3 sequences were included. The clonal type of each cell was determined by their nucleotide sequences of CDR3 region in both alpha/beta or gamma/delta chains. Clonal types appeared in more than 1 cells in each sample were considered as clonal expansion, otherwise considered as unique. The SARS-CoV2 and common viruses' specificities were obtained by applying the GLIPH2 algorithm[6]. In brief, the VDJ sequencing results of $CD8^+$ T cells were GLIPH2 clustered with public datasets of SARS-CoV2-specific or common viruses-specific TCR sequences, which were generated by experimental approaches. TCRs that were clustered with the TCRs of known specificity in the public datasets were labelled accordingly. The GLIPH2 clustering is generally based on the CDR3 sequence similarity with different weights at different positions according to the crystallization results of TCR-peptide-MHC binding.

**Table 1. Broad extension panel of antibodies used in mass cytometry**

| Metal tag | Marker | Clone | Vendor* |
|---|---|---|---|
| 89Y | CD45 | HI30 | Standard BioTools |
| 102Pd | Barcode | - | Standard BioTools |
| 104Pd | Barcode | - | Standard BioTools |
| 105Pd | Barcode | - | Standard BioTools |
| 106Pd | Barcode | - | Standard BioTools |
| 108Pd | Barcode | - | Standard BioTools |
| 110Cd | CD33 | WM53 | Biolegend |
| 111Cd | CD26 | BA5b | Biolegend |
| 112Cd | CD11c | Bu15 | Biolegend |
| 113Cd | IgD | IA6-2 | Biolegend |
| 114Cd | HLA-DR | L243 | BioLegend |
| 115In | CD57 | HCD57 | Biolegend |
| 140Ce | CD71 | CY1G4 | Biolegend |
| 141Pr | CD49d | 9F10 | Standard BioTools |
| 142Nd | CD43 | 84-3C1 | eBiosciences |
| 143Nd | CD3e | UCHT1 | Biolegend |
| 144Nd | CD15 | W6D3 | Biolegend |
| 145Nd | CD81 | 5A6 | Biolegend |
| 146Nd | CD52 | HI186 | Biolegend |
| 147Sm | CD1c | L161 | Biolegend |
| 148Nd | CD55 | JS11 | Biolegend |
| 149Sm | CD25 | 2A3 | Standard BioTools |
| 150Nd | CD64 | 10.1 | Biolegend |
| 151Eu | CD123 | 6H6 | BioLegend |
| 152Sm | TCRgd | 5A6.E9 | Thermo Fisher Scientific |
| 153Eu | Siglec-8 | 837535 | R&D Systems |
| 154Sm | CD95 | DX2 | BioLegend |
| 155Gd | CX3CR1 | 8E10.D9 | BioLegend |
| 156Gd | CD20 | 2H7 | BioLegend |
| 157Gd | CD9 | SN4 C3-3A2 | eBiosciences |
| 158Gd | KIR2DL2/L3 | DX27 | Biolegend |
| 158Gd | KIR2DL1/KIR2DS5 | 143211 | R&D |
| 158Gd | KIR3DL2/CD158k | 539304 | R&D |
| 158Gd | KIR3DL1 | DX9 | BD |
| 158Gd | KIR2DL5 | UP-R1 | Miltenyi |
| 159Tb | CD22 | HIB22 | Biolegend |
| 160Gd | CD14 | M5E2 | Biolegend |
| 161Dy | CD161 | HP-3G10 | Biolegend |

| | | | |
|---|---|---|---|
| 162Dy | CD29 | TS2/16 | Biolegend |
| 163Dy | 4-1BB | 4B4-1 | Biolegend |
| 164Dy | CD10 | HI10a | Biolegend |
| 165Ho | CD127 | A019D5 | Standard BioTools |
| 166Er | CD24 | ML5 | Biolegend |
| 167Er | CD27 | L128 | Biolegend |
| 168Er | CD141 | M80 | Biolegend |
| 169Tm | CD45RA | HI100 | Standard BioTools |
| 170Er | CD38 | HIT2 | Biolegend |
| 171Yb | CD85j | GHI/75 | Biolegend |
| 172Yb | CD279 (PD-1) | EH12.2H7 | BioLegend |
| 173Yb | CD56 | NCAM16.2 | BD Biosciences |
| 174Yb | CD99 | HCD99 | Biolegend |
| 175Lu | CD28 | CD28.2 | Biolegend |
| 176Yb | CD39 | A1 | Biolegend |
| 191Ir | DNA Ir | Cell-ID DNA Intercalator | Standard BioTools |
| 193Ir | DNA Ir | Cell-ID DNA Intercalator | Standard BioTools |
| 194Pt | CD8a | SK1 | BD Biosciences |
| 195Pt | CD5 | UCHT2 | Biolegend |
| 196Pt | CD7 | CD7-6B7 | Biolegend |
| 198Pt | CD4 | RPA-T4 | Biolegend |
| 209Bi | CD16 | 3G8 | Standard BioTools |

**All antibodies that are not from Standard BioTools were purchased in purified format and coupled in-house*

**Table 2. Antibodies used in memory T cell isolation**

| Cat # | Name |
|---|---|
| 301826 | Biotin Anti human CD14 |
| 301914 | Biotin Anti human CD15 |
| 302004 | Biotin Anti human CD16 |
| 302204 | Biotin Anti human CD19 |
| 343524 | Biotin Anti human CD34 |
| 336218 | Biotin Anti human CD36 |
| 304104 | Biotin Anti human CD45RA |
| 304412 | Biotin Anti human CD51/61 |
| 306004 | Biotin anti human CD123 |
| 344108 | Biotin anti human CD141 |
| 306618 | Biotin anti human CD235ab |

#### *SARS-CoV-2 plasma antigen analyses using SIMOA*

SARS-CoV-2 spike, S1, and nucleocapsid antigens were measured in plasma samples as previously described[7]. Briefly, samples were thawed and centrifuged at 2000 x g for

10 minutes at 4 °C to remove cellular debris. The samples were then diluted with a solution of 10 mM dithiothreitol (DTT), protease inhibitors, and incubated at 37 °C allowing the DTT to denature any antibodies bound to circulating antigens. Using an HD-X Analyzer (Quanterix Corporation, Billerica MA), 100 µL of diluted plasma was incubated with beads coated with anti-SARS-CoV-2 antibodies. After washing, the beads were resuspended in detector antibody solution, incubated and washed. Next, the beads were resuspended in streptavidin conjugated β-galactosidase solution, incubated, and washed before being resuspended in resorufin β-D-galactopyranoside and loaded into a microwell array for imaging.

**COVID Human Genetic Effort consortium**

The members of the COVID Human Genetic Effort are as follows: Laurent Abel (Laboratory of Human Genetics of Infectious Diseases, Necker Branch, INSERM U1163, Necker Hospital for Sick Children, Paris, France; Paris Cite University, Imagine Institute, Paris, France; and St. Giles Laboratory of Human Genetics of Infectious Diseases, Rockefeller Branch, Rockefeller University, New York, NY), Salah Al-Muhsen (Immunology Research Lab, Department of Pediatrics, College of Medicine, King Saud University, Riyadh, Saudi Arabia), Fahd Al-Mulla (Dasman Diabetes Institute, Department of Genetics and Bioinformatics, Dasman, Kuwait), Evangelos Andreakos (Laboratory of Immunobiology, Center for Clinical, Experimental Surgery and Translational Research, Biomedical Research Foundation of the Academy of Athens, Athens, Greece), Novelli Antonio (Laboratory of Medi-cal Genetics, IRCCS Bambino Gesu Children's Hospital, Rome, Italy), Andres A. Arias (Group of Primary Immunodeficiencies, University of Antioquia Ude A, Medellin, Colombia), Sophie Trouillet-Assant (Hospices Civils de Lyon, International Center of Research in Infectiology, Lyon University, INSERM U1111, CNRS UMR 5308, ENS, UCBL, Lyon, France), Alexandre Belot (Pediatric Nephrology, Rheumatology, Dermatology, HFME, Hospices Civils de Lyon, National Referee Centre RAISE, and INSERM U1111, Université de Lyon, Lyon, France), Catherine M. Biggs (Department of Pediatrics, British Columbia Children's Hospital, The University of British Columbia, Vancouver, British Columbia,

Canada), Ahmed A. Bousfiha (Clinical Immunology Unit, Department of Pediatric Infectious Disease, CHU Ibn Rushd and the Laboratoire d'Immunologie Clinique, Inflammation et Allergie, Faculty of Medicine and Pharmacy, Hassan II University, Casablanca, Morocco), Alexandre Bolze (Helix, San Mateo, Calif), Alessandro Borghesi (Fondazione IRCCS Policlinico San Matteo di Pavia, Fellay Laboratory, Ecole Polytechnique Federale de Lausanne, Lausanne, Switzerland), John Christodoulou (Murdoch Children's Research Institute and Department of Paediatrics, University of Melbourne, Melbourne, Australia), Aurelie Cobat (Laboratory of Human Genetics of Infectious Diseases, Necker Branch, INSERM U1163, Necker Hospital for Sick Children, Paris, France; Paris Cité University, Imagine Institute, Paris, France; and St. Giles Laboratory of Human Genetics of Infectious Diseases, Rockefeller Branch, Rockefeller University, New York, NY), Antonio Condino-Neto (Department of Immunology, Institute of Biomedical Sciences, University of Sao Paulo, Sao Paulo, Brazil), Stefan Constantinescu (de Duve Institute and Ludwig Cancer Research, Brussels, Belgium), Clifton L. Dalgard (Department of Anatomy, Physiology and Genetics, Uniformed Services University of the Health Sciences, Bethesda, Md), Xavier Marie Duval (INSERMCIC 1425 Paris, France; AP-HP, University Hospital of Bichat Paris, France, University Paris Diderot, Paris 7, UFR de Medecine-Bichat, Paris, France; IAME, INSERM, UMRS1137, University of Paris, Paris, France; and AP-HP, Bichat Claude Bernard Hospital, Infectious and Tropical Diseases Department, Paris, France), Sara Espinosa-Padilla (National Institute of Pediatrics, Mexico City, Mexico), Jacques Fellay (School of Life Sciences, Ecole Poly-technique Federale de Lausanne, Lausanne, Switzerland, and Precision Medicine Unit, Lausanne University Hospital and University of Lausanne, Lausanne, Switzerland), Carlos Flores (Genomics Division, Instituto Tecnologico y de Energıas Renovables, Santa Cruz de Tenerife, Spain; Research Unit, Hospital Universitario N.S. de Candelaria, Santa Cruz de Tenerife, Spain; CIBER de Enfermedades Respiratorias, Instituto de Salud Carlos III, Madrid, Spain; Department of Clinical Sciences, University Fernando Pessoa Canarias, Las Palmas de Gran Canaria, Spain), José Luis Franco (Group of Primary Immunodeficiencies, University of Antioquia Ude A, Medellin, Colombia), Antoine Froidure (Pulmonology Department, Cliniques Universi-taires Saint-Luc, and Institut de

Recherche Experimentale et Clinique), Université Catholique de Louvain, Brussels, Belgium), Guy Gorochov (Sorbonne Université, Inserm, Centre d'Immunologie et des Maladies Infectieuses Paris, Assistance Publique-Hopitaux de Paris (AP-HP) HopitalPitie-Salpetriere, Paris, France), Filomeen Haerynck (Department of Paediatric Immunology and Pulmonology, Centre for Primary Immunodeficiency, Ghent, PID Research Laboratory, Jeffrey Modell Diagnosis and Research Centre, Ghent University Hospital, Ghent, Belgium), Rabih Halwani (Sharjah Institute of Medical Research, College of Medicine, University of Sharjah, Sharjah, United Arab Emirates), Elena W. Y. Hsieh (Departments of Pediatrics, Immunology and Microbiology, University of Colorado, School of Medicine, Aurora, Colorado), Yuval Itan (Institute for Personalized Medicine, Icahn School of Medicine at Mount Sinai, NY, and Department of Genetics and Genomic Sciences, Icahn School of Medicine at Mount Sinai, New York, NY), Erich D. Jarvis (Laboratory of Neurogenetics of Language, HHMI, The Rockefeller University, New York, NY), Kai Kisand (Molecular Pathology, Department of Biomedicine, Institute of Biomedicine and Translational Medicine, University of Tartu, Tartu Estonia), Yu-Lung Lau (Department of Paediatrics and Adolescent Medicine, The University of Hong Kong, Hong Kong, China), Davood Mansouri (Department of Clinical Immunology and Infectious Diseases, National Research Institute of Tuberculosis and Lung Diseases, The Clinical Tuberculosis and Epidemiology Research Center, National Research Institute of Tuberculosis and Lung Diseases, Masih Daneshvari Hospital, Shahid Beheshti, University of Medical Sciences, Tehran, Iran), Trine H. Mogensen (Department of Biomedicine, Aarhus University, Aarhus, Denmark), Lisa F. P. Ng (A*STAR Infectious Disease Labs, Agency for Science, Technology and Research, Singapore, and Lee Kong Chian School of Medicine, Nanyang Technology University, Singapore), Luigi D. Notarangelo (National Institute of Allergy and Infectious Diseases, National Institutes of Health, Bethesda, MD), Giuseppe Novelli (Department of Biomedicine and Prevention, Tor Vergata University of Rome, Rome, Italy), Satoshi Okada (Department of Pediatrics, Graduate School of Biomedical and Health Sciences, Hiroshima University, Hiroshima, Japan), Tayfun Ozcelik (Department of Molecular Biology and Genetics, Bilkent University, Bilkent, Ankara, Turkey), Rebeca Perez de Diego

(Laboratory of Immunogenetics of Human Diseases, Innate Immunity Group, IdiPAZ Institute for Health Research, La Paz Hospital, Madrid, Spain), Carolina Prando (Faculdades Pequeno Prıncipe, Instituto de Pesquisa Pele Pequeno Prıncipe, Curitiba, Brazil), Aurora Pujol (NeurometabolicDiseases Laboratory, Bellvitge Biomedical Research Institute, L'Hospitalet de Llobregat, Barcelona, Spain; Catalan Institution of Research and Advanced Studies, Barcelona, Spain, and Center for Biomedical Research on Rare Dis-eases, ISCIII, Barcelona, Spain), Lluis Quintana-Murci (Human Evolutionary Genetics Unit, CNRS U2000, Institut Pasteur, Paris, France, and Human Ge-nomics and Evolution, College de France, Paris, France), Laurent Renia (A*STAR Infectious Disease Labs, Agency for Science, Technology and Research, Singapore; Lee Kong Chian School of Medicine, Nanyang Technology University, Singapore), Igor Resnick (Department of Medical Genetics, Medical University, Varna and Department Hematology and BMT, University Hospital St. Marina, Bulgaria), Lucie Roussel (Department of Medicine, Di-vision of Infectious Diseases, McGill University Health Centre, Montreal, Quebec, Canada, and Infectious Disease Susceptibility Program, Research Institute, McGill University Health Centre, Montreal, Quebec, Canada), Carlos Rodrıguez-Gallego (Department of Immunology, University Hospital of Gran Canaria Dr. Negrın, Canarian Health System, Las Palmas de Gran Cana-ria, Spain; Department of Medical and Surgical Sciences, School of Medicine, University of Las Palmas de Gran Canaria, Las Palmas de Gran Canaria, Spain, and Department of Clinical Sciences, University Fernando Pessoa Canarias, Las Palmas de Gran Canaria, Spain), Vanessa Sancho-Shimizu (Department of Paediatric Infectious Diseases and Virology, Imperial College London, London, UK, and Centre for Paediatrics and Child Health, Faculty of Medicine, Imperial College London, London, UK), Mohammad Shahrooei (Specialized Immunology Laboratory, Ahvaz, Iran; Department of Microbiology and Immunology, Clinical and Diagnostic Immunology, Katholieke Universiteit Leuven, Leuven, Belgium), Pere Soler-Palacın (Pediatric Infectious Diseases and Immunodeficiencies Unit, Vall d'Hebron Barcelona Hospital Campus, Barcelona, Spain), Ivan Tancevski (Department of Internal Medicine II, Medical University of Innsbruck, Innsbruck, Austria), Stuart G. Tangye (Garvan Institute of Medical Research, Darlinghurst, New South Wales,

Australia, and St. Vincent's Clinical School, Faculty of Medicine, University of New South Wales, Sydney, New South Wales, Australia), Ahmad Abou Tayoun (Al Jalila Children's Hospital, Dubai,United Arab Emerates), Sehime Gülsün Temel (Department of Medical Genetics, Department of Histology and Embryology, Faculty of Medicine, Bursa Uludag University, Bursa, Turkey; and Department of Translational Medicine, Health Sciences Institute, Bursa Uludag University, Bursa, Turkey), Pierre Tiberghien (Etablissement Francais Du Sang, La Plaine-Saint Denis, Saint-Denis, France) Jordi Perez Tur (Institut de Biomedicina de Valencia-CSIC, CI-BERNED, Unitat Mixta de Neurologia i Genetica, IIS La Fe, Vallencia, Spain), Stuart E. Turvey (BC Children's Hospital, The University of British Columbia, Vancouver, Canada), Furkan Uddin (Centre for Precision Therapeutics, Genetics and Genomic Medicine Centre, Holy Family Red Crescent Medical College Dhaka, Bangladesh) Mohammed J. Uddin (College of Medicine, Mohammed Bin Rashid University of Medicine and Health Sciences, Dubai, United Arab Emirates; Cellular Intelligence Lab, Genome Arc, Inc, Toronto, Ontario, Canada), Mateus Vidigal (University of Sao Paulo, Brazil), Donald C. Vinh (Department of Medicine, Division of Infectious Diseases, McGill University Health Centre, Montreal, Quebec, Canada, and Infectious Disease Susceptibility Program, Research Institute, McGill University Health Centre, Montreal, Quebec, Canada), Mayana Zatz (Biosciences Institute, University of Sao Paulo, Sao Paulo, Brazil), Keisuke Okamoto (Tokyo Medical and Dental University, Tokyo, Japan), David S. Perlin (Center for Discovery and Innovation, Hackensack Meridian Health, Nutley, NJ), Graziano Pesole (Department of Biosciences, Biotechnology and Biopharmaceutics, University of Bari A. Moro, Bari, Italy), Christian Thorball (Precision Medicine Unit, Lausanne University Hospital and University of Lausanne, Lausanne, Switzerland.) Diederik van de Beek (Department of Neurology, Amsterdam Neuroscience, Amsterdam University Medical Center, University of Amsterdam, Amsterdam, The Netherlands), Roger Colobran (Hospital Universitari Vall d'Hebron, Barcelona, Spain), Joost Wauters (Department of General Internal Medicine, Medical Intensive Care Unit, University Hospitals Leuven, Leuven, Belgium), Shen-Ying Zhang (Laboratory of Human Genetics of Infectious Diseases, Necker Branch, INSERM U1163, Necker Hospital for Sick Children, Paris, France; Paris Cite University, Imagine

Institute, Paris, France; and St. Giles Laboratory of Human Genetics of Infectious Diseases, Rockefeller Branch, Rockefeller University, New York, NY), Qian Zhang (Laboratory of Human Genetics of Infectious Diseases, Necker Branch, INSERMU1163, Necker Hospital for Sick Children, Paris, France; Paris Cite University, Imagine Institute, Paris, France; and St. Giles Laboratory of Human Genetics of Infectious Diseases, Rockefeller Branch, Rockefeller University, New York, NY), Helen C. Su (National Institute of Allergy and Infectious Diseases, National Institutes of Health, Bethesda, MD).
